# Preventing within household transmission of COVID-19: Is the provision of accommodation feasible and acceptable?

**DOI:** 10.1101/2020.08.20.20176529

**Authors:** Sarah Denford, Kate Morton, Jeremy Horwood, Rachel de Garang, Lucy Yardley

**Author notes:** Correspondence to: Dr Sarah Denford, Population Health Sciences, Bristol Medical School, University of Bristol, Priory Road Complex, Bristol, BS8 1TU.

## Abstract

**Background:** Within-household transmission of COVID-19 is responsible for a significant number of infections. The risk of within-household infection is greatly increased among those from Black Asian and minority ethnic (BAME) and low income communities. Efforts to protect these communities are urgently needed. The aim of this study is to explore the acceptability of the availability of accommodation to support isolation among at risk populations.

**Methods:** Our study used a mixed methods design structured in two phases. In phase 1, we conducted a survey study of a sample of volunteers from our existing database of 300 individuals who had provided consent to be contacted about ongoing research projects into infection control. In phase 2, we conducted semi-structured interviews with 19 participants from BAME communities and low income communities recruited through social media.

**Results:** Participants from the survey and interview phase of the study viewed the provision of accommodation as important and necessary. Factors influencing likely uptake of accommodation included perceived 1) vulnerability of household 2) exposure to the virus and 3) options for isolation at home. Barriers to accepting the offer of accommodation included 1) being able to isolate at home 2) wanting to be with family 3) caring responsibilities 4) concerns about mental wellbeing 5) upheaval of moving when ill and 6) concerns about infection control. Participants raised a series of issues that should be addressed before accommodation is offered. These included questions regarding who should use temporary accommodation and at what stage to effectively reduce transmission in the home, and how infection control in temporary accommodation would be managed.

**Conclusion:** This research provides evidence that the provision of accommodation to prevent within household transmission of the virus is viewed as acceptable, feasible and necessary by many people who are concerned about infection transmission in the home. We explore ways in which accommodation might be offered. In particular, vulnerable members of the household could be protected if accommodation is offered to individuals who are informed through test trace and isolate that they have been in contact with the virus.

## Introduction

Human behaviour is central to the transmission of COVID-19. To reduce transmission in the absence of pharmaceutical interventions, a series of behavioural interventions have been suggested and implemented (1). Whilst the introduction of social distancing behaviours can reduce the spread of COVID-19 within the community (2), people with symptoms of the virus are instructed to remain in the home; potentially with cohabiting families and friends. This has led to clusters of infection within households (3), and within household transmission being highlighted as a dominant route of infection (4, 5). In order to avoid within household transmission of COVID-19, excellent infection control measures are needed (4). This includes introducing hygiene protocols, appropriate use of personal protective equipment (e.g., face mask use when necessary), and within household distancing and segregation – or ‘self-isolation’ – of infected individuals (5-7). Although effective for reducing within household transmission (2, 8), there is substantial variation in the extent to which the public are able and willing to adhere to these behavioural solutions (9-11).

There is little doubt that COVID-19 is exposing and widening existing inequalities within society. Data have shown that those from Black, Asian, and minority ethnic (BAME) communities have a markedly higher risk of infection (12, 13) and worse clinical outcomes, including intensive therapy unit (ITU) admission and mortality (14, 15). Likewise, those living in the most deprived areas are more likely to be diagnosed with COVID-19, and have worse outcomes than those living in the least deprived areas (15). The reasons underpinning the disproportionate impact of the virus on these populations are multiple and complex, but include increased risk of occupational and geographical exposure (12, 15-17) paired with reduced opportunities for social distancing and self-isolation (9). Indeed, a recent survey highlighted that the ability to isolate was lowest among low income households and those from BAME communities (9). People from these communities were also considerably less likely to be able to work remotely. Innovative solutions to prevent the spread of the virus within BAME and low income group households are therefore urgently required.

One potential solution to preventing the transmission of the virus within the home is isolation outside the home. Centralised – as opposed to individual – isolation has been suggested (3) and implemented successfully in locations such as China and Korea (18). In Wuhan, for example, existing public venues were rapidly converted into what are termed ‘Fangcang shelter hospitals’. Individuals with symptoms of COVID-19 would isolate within these shelters, away from friends and family. In addition to providing food and medical care, these locations ensured adherence to self-isolation guidance, keeping the families and household members of the infected individual safe from infection, and provided social engagement, reducing psychological distress associated with self-isolation (19). Indeed, a key difference between Fangcang shelters and makeshift or emergency hospitals is the social space provided, allowing residents to engage and socialise with others during the isolation period (19). However, although cost effective and acceptable to residents living in Wuhan, the substantial differences in culture and living conditions mean that Fangcang style accommodation may be less likely to be accepted by individuals in many European countries.

A small number of European countries have however, converted some hotels, hostels, dormitories or specialised facilities into special facilities to accommodate people who are experiencing symptoms of the virus (20). This strategy has not been widely implemented, and most of Europe and the United States continue to encourage individuals with symptoms to self-isolate within the home. In the United Kingdom, National Health Service (NHS) workers were offered, on a voluntary basis, the option of staying in NHS reimbursed hotel accommodation to enable them to continue to work if they were living with others who may be vulnerable. For those who can afford it, self-funded luxury hotel “quarantine packages” are available (21-23), but, funded accommodation has not yet been offered in the UK to individuals or communities outside the NHS, who may be at risk.

Whilst the offer of funded accommodation within which to self-isolate or quarantine is a potentially viable strategy, it is critical that interventions are culturally appropriate and acceptable to the communities that they serve to protect. This requires extensive input from target users to understand the environmental and cultural context within which the intervention could be introduced, as well as the psychological and social factors likely to influence uptake (9).

The aim of this research is therefore to understand whether or not offers of accommodation would be acceptable and feasible for people concerned about reducing infection transmission in the home, and among BAME and low income communities, and to elicit discussions regarding what we can do to improve advice and approaches to reduce transmission of the virus within the home.

## Methods

### Study design

Our study used a mixed methods design structured in two phases. In phase 1, we conducted a survey study of a sample of volunteers from our existing database of 300 individuals who had been recruited through their engagement with Germ Defence, a website aiming to reduce infection risks in the home (Supplement 1). These individuals had provided consent to be contacted about ongoing research projects. In phase 2, we conducted semi-structured interviews with 19 participants from BAME and low income communities. These interviews were designed to help us explore further concepts identified in phase 1, and to elicit discussions regarding how accommodation could best be utilised.

### Phase 1

#### Data collection

A convenience sample of volunteers who had previously provided consent to be contacted and invited to take part in research were recruited via email distribution lists. Participants were invited to complete a confidential online survey regarding their ability and willingness to isolate within the home, and the acceptability of accommodation to isolate outside the home. Informed consent was collected online before starting the survey.

#### Data analysis

Frequencies and descriptive statistics are presented for closed survey questions. Free text answers were used to offer further insight into, and explanations for, answers given to closed survey questions. We identified barriers and facilitators related to the provision of accommodation with qualitative content analysis in three stages (24, 25). First, responses to the survey were coded by two authors independently. During stage two, codes were categorised into a unique list of barriers and facilitators, which were discussed and refined by the same two authors. Data were then assigned to each category, and counts of text assigned to each category were generated.

### Phase 2

#### Data collection

Volunteers from BAME and low income communities were recruited via existing contacts with community groups, social media advertisements, and snowball sampling. Interested individuals responded to an invitation take part in research to understand experiences and interpretations of self-isolation and protection during the pandemic. Participants were over the age of 18 years and residing in the UK. We purposely sampled for diversity in key factors, including ethnicity, living arrangements, occupation, and vulnerability. Sample size was informed by the concept of ‘information power’, (26) with analysis and sampling conducted in parallel and continuous assessment of the suitability of the information within the sample with regard to study objectives. Potential participants contacting the research team were provided with a study information sheet and given an opportunity to ask any questions. Participants were informed of the voluntary nature of participation in the study, and assured of the confidentiality of the data collected. As all interviews were conducted via the telephone or online, audio recorded (rather than written) verbal consent was obtained.

The semi-structured topic guide (Supplement 2) was informed by data collected during phase 1 as well as existing literature, and conversations with experts in public health, behaviour change and intervention development. Questions were designed to explore participants’ current living situation, their experiences of isolation within the home, perceptions relating to the provision of accommodation to reduce transmission in the home, and suggestions regarding how accommodation may be used and facilitated. Interviews lasted between 21 and 55 minutes (mean duration 38 minutes).

#### Analysis

Data from the interviews were analysed using a thematic approach aimed at identifying issues raised by the participants and ways in which these issues may be mitigated (27). Following the stages of thematic analysis, two researchers independently read transcripts to assign codes to the data and identify possible themes. These themes were discussed and refined through discussion. An initial framework was developed, checked against the data, and refined as necessary. Charts were developed for each theme in the framework, and relevant text from the transcripts were be copied or summarised under each theme in the chart. Charts were then used to compare data within and between individuals. Participants were invited to discuss the analysis and interpretations with the researchers via skype or email.

## Results

### Phase 1

A total of 110 respondents completed the survey (Table 1). Of all respondents, 24 (22%) stated that they would accept an offer of accommodation if it was available, 25 (22%) said that they would probably accept, 21 (19%) said they would probably not accept and 39 (35%) said that they would not accept. Of the 85 (77%) participants who said they were not able to isolate at home, 24 (28%) said they would accept, 23 (27%) said that they would probably accept, 18 (21%) said that they would probably not accept, and 16 (18%) said they would not accept. Of those unable to isolate at home, and who also considered themselves to be of high risk if they catch the virus (N = 36) or living with someone who is high risk (N 18), a total of 19 (35%) said that they would accept, 12 (22%) would probably accept, 14 (26%) would probably not accept, and 8 (14%) would not accept.

**Table 1:** Participant characteristics

|  | Phase 1 (N=110) | Phase 2 (N=19) |
| --- | --- | --- |
| **Age** |  |  |
| 18-25 | 0 | 2 |
| 26-40 | 2 | 8 |
| 41-60 | 35 | 8 |
| 61-70 | 41 | 1 |
| Over 70 | 30 | 0 |
| Missing | 2 | 0 |
| **Gender** |  |  |
| Male | Missing | 7 |
| Female | Missing | 12 |
| **Ethnic group** |  |  |
| White | 104 | 6 |
| Mixed / multiple ethnic groups | 1 | 1 |
| Asian / Asian British | 0 | 9 |
| Black / African / Caribbean / Black British | 1 | 3 |
| Missing | 4 | 0 |
| **Leaving full time education** |  |  |
| Before finishing school | 1 | 1 |
| After finishing school | 42 | 4 |
| After finishing university | 36 | 4 |
| After postgraduate studies | 28 | 1 |
| Missing | 3 | 9 |
| **Experience with COVID-19** |  |  |
| I am at increased risk | 48 | 1 |
| I live with someone high risk | 19 | 7 |
| I have had the virus | 7 | 1 |
| I live with someone who had the virus | 1 | 0 |
| None of the above | 32 | 10 |
| Missing | 3 | 0 |

Three factors were coded as facilitators influencing decisions to accept an offer of accommodation to reduce transmission in the home (Table 2). These were to protect others within the household, to control the virus, and to avoid using shared spaces. Seven barriers to accepting the offer of accommodation included 1) the ability to isolate within the home, 2) not wanting to be apart from family 3) having caring responsibilities (4) concerns about the impact of isolation on mental wellbeing and relationships (5) concerns about the upheaval of moving when ill, (6) perceived risk of catching or spreading coronavirus if leaving the building, and (7) unfeasible for unspecified reasons (Table 2).

**Table 2:**
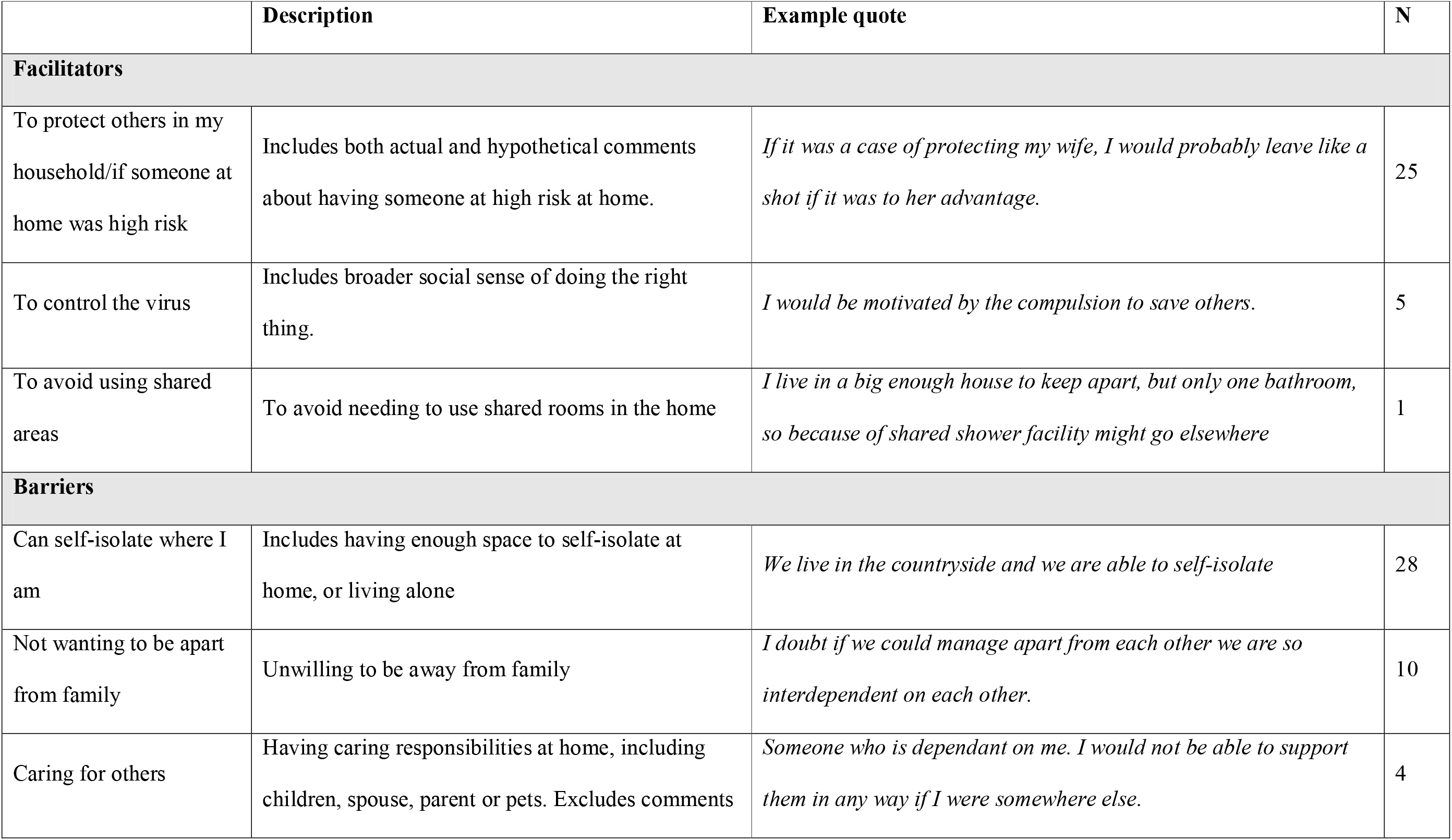
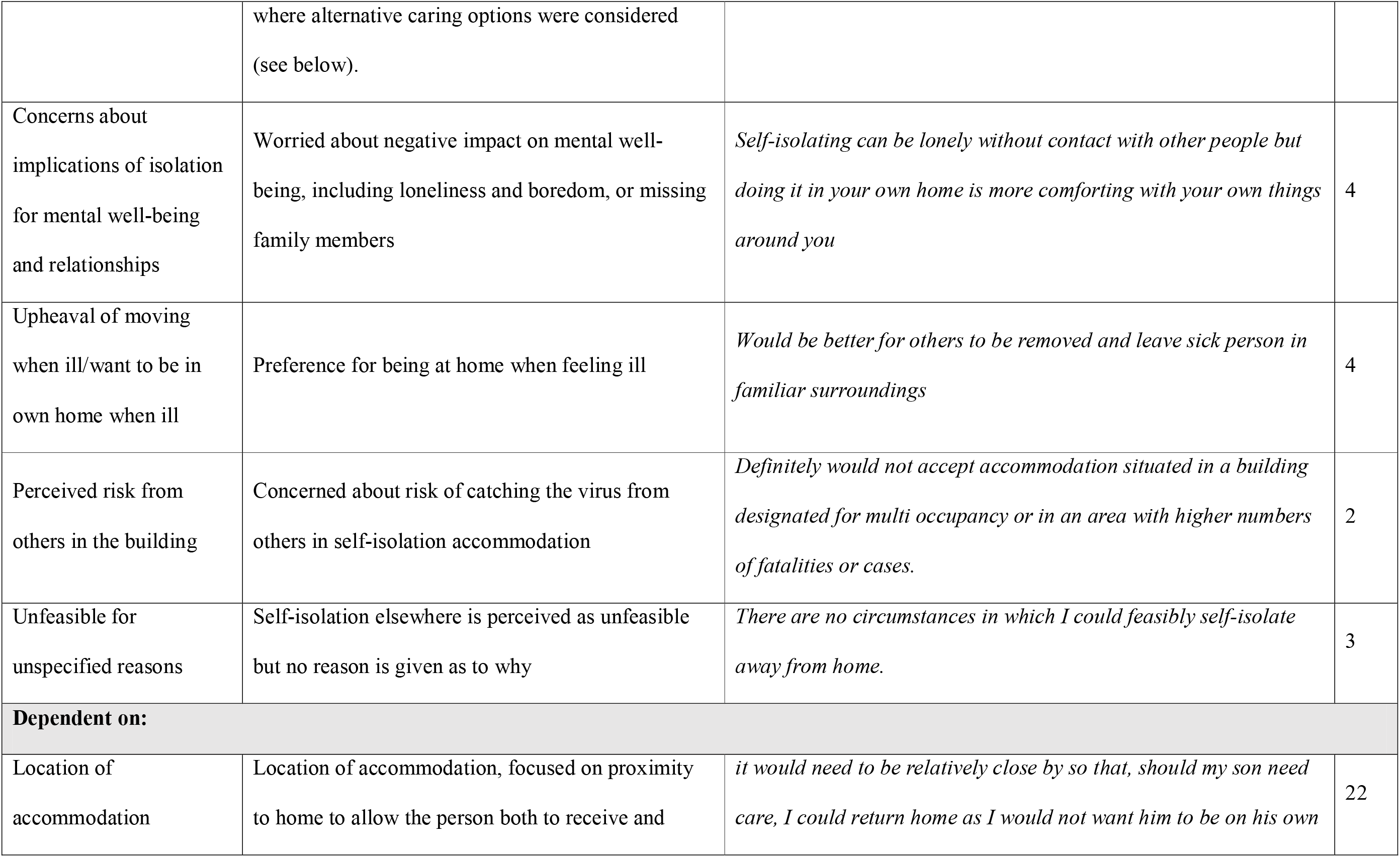
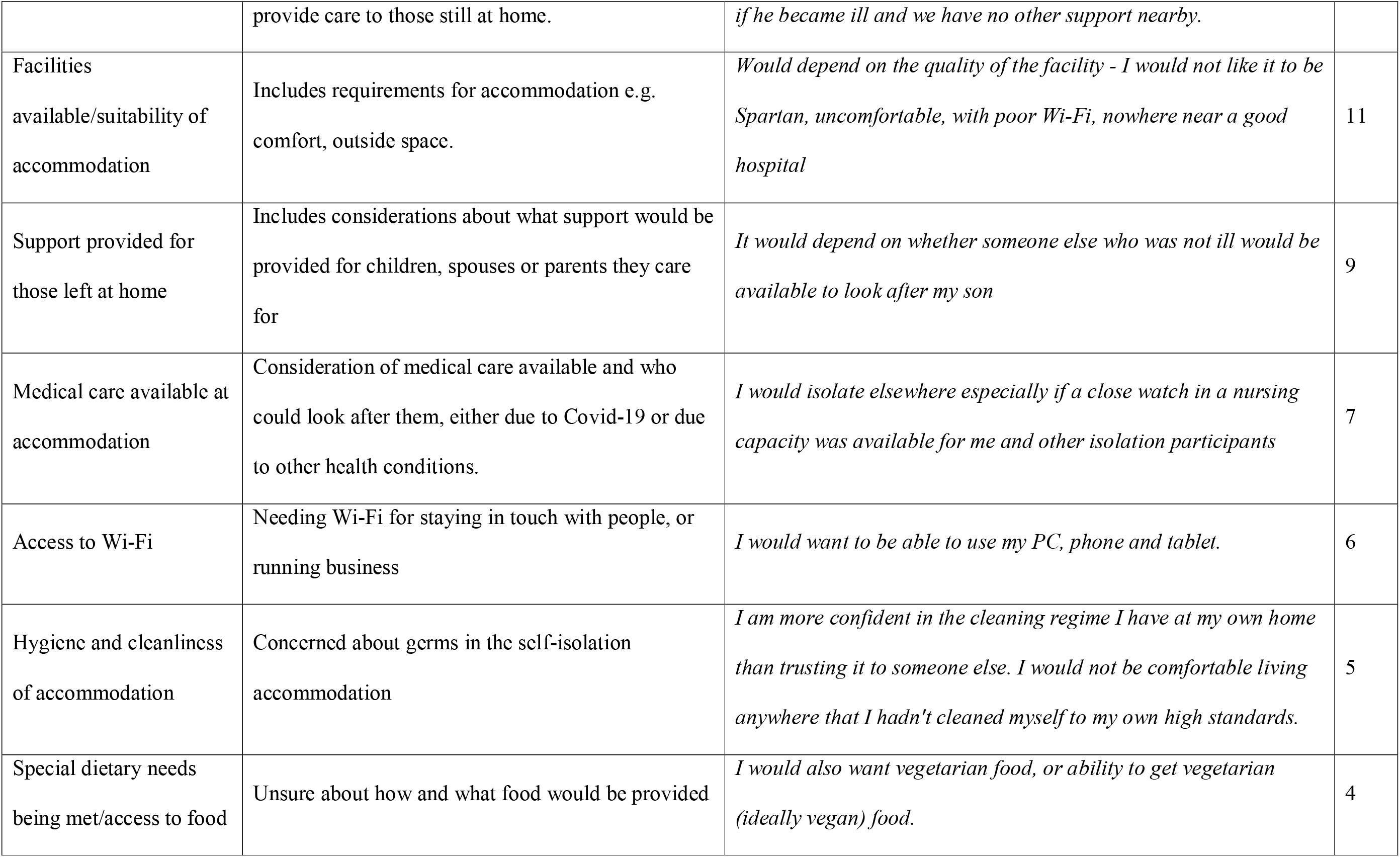
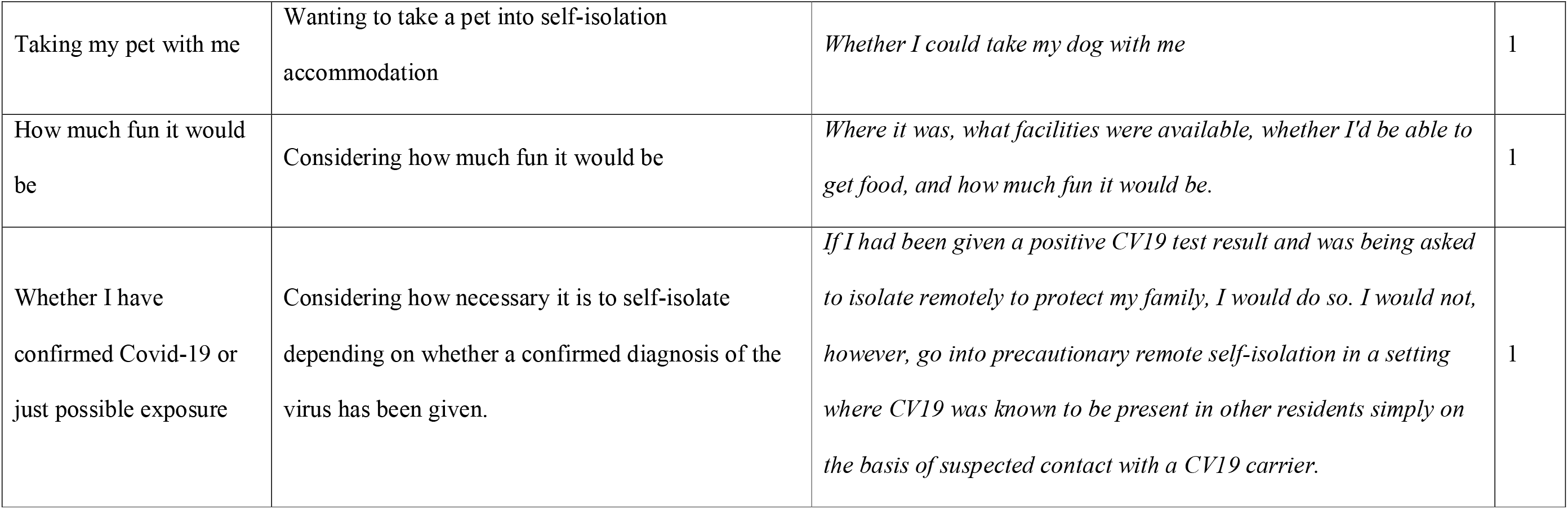
Facilitators and barriers to the uptake of accommodation for isolation – results of the content analysis of survey text

### Phase 2

A total of 19 participants took part in the interviews from Black African (N=2), Black British (N=1), Mixed White / Black Caribbean (N=1) Indian (N=5), British Indian (N=2) Asian (N=1) British Asian Pakistani (N=1) and White (N=6) ethnic groups (Table 1). Only two participants reported that they would be unlikely to accept the offer of accommodation outside the home. Six participants said that they would accept as a last resort, four said that they would have accepted the offer had it been needed, and six participants said that they would be likely to accept. One participant had moved a family member out of the home for 11 weeks during the pandemic.

### Protecting the household

Participants were positive about the idea of accommodation being offered to reduce transmission of the virus in the home. It was considered to be a highly effective way of preventing the spread of the virus among those who were unable to isolate within their current homes.

*“If I was offered accommodation which meant that my family were kept safe, then absolutely I would, I would welcome it” (Participant 14, White, female)*.

Critically, participants thought that it had the potential to save lives:

*“Wow that would probably have saved a lot of lives actually. Yeah” (Participant 03, British Asian Pakistani, male)*.

### Risk

The decision to accept, or not, the offer of accommodation appeared to be influenced by how at risk the person considered themselves or their household to be. Perceived risk was influenced by how vulnerable the participant (or their household) were perceived to be, level of exposure to the virus, and level of contact with household members.

### Vulnerability

Eight participants considered themselves or a member of their household to be vulnerable, and this was strongly influential in decisions regarding the use of temporary accommodation. One participant, whose husband had moved out of family home for 11 weeks over the pandemic, explained how keeping her vulnerable daughter safe was their main priority:

“*It’s just something that has to be done, you know, and he actually didn’t come back inside the house, he left for work that morning and then didn’t come back for 11 weeks. His bags were packed, his bags were packed and the hotel was booked by the evening and gone” (Participant 18, White, female)*.

Participants who did not consider themselves (or their household) to be vulnerable reported that they would be more willing to accept the offer of accommodation outside the home if they or their family were vulnerable:

*“Yeah maybe if I had my older relatives with me, or I had somebody who um, you know had any underlying health condition, probably yeah I would have offered to go out, but in the current situation I wouldn’t have, so. If I had somebody who was living with me who was over 65 years old or who had heart disease or was diabetic, I would offered to go out yes of the house” (Participant 06, Indian, female)*.

Due to the severity of the virus, any one could consider themselves to be vulnerable, regardless of age and health status:

*“I even read on the net or so, I’m not sure if this information is credible or not, but still what I saw on the net is even if you get the virus even if you recover from it, it can have detrimental consequences on your health. For example I read somewhere on the net I read that if you have the virus it can damage your lungs, like, forever, it can have impact on your lungs forever, so this bit of information is quite scary” (Participant 07, Indian, female)*.

### Exposure to the virus

Accommodation was considered to be particularly important for those who are in situations in which there is potential for high exposure to the virus. There was wide understanding that those from BAME and low income communities were more likely to be in situations in which exposure to the virus is probable:

*“Lots of people of colour, and not just, Bangladeshi etcetera, who work in jobs where they have no choice but to go in. You know, if someone said isolate, they would say ‘well how will I feed my family?’ They have to go in. So they’re in jobs where they have to go in, they have to mix with the public” (Participant 11, Black African, female)*.

Participants even described situations in which people from BAME communities were asked to leave accommodation due to increased exposure to the virus:

*“ When he got back [from work] the door locks were changed and she [the landlady] said ‘I’m really sorry but I can’t have you in here because I’m too frightened, you’re a cab driver, you’re seeing all these people you’re going to infect the whole house you know, I’m sorry I can’t have you in here’” (Participant 11, Black African, female)*.

Whilst recognising the value of accommodation, those who were not exposed to the virus thought that accommodation would be unnecessary for their household:

*“If I’m not taking the precaution for example, if I have to go to work, then yeah I would suggest for him to isolate somewhere else because I might have the virus in transfer it to him, so yeah. But my case is different because I work from home and I’m not going out and I’m not meeting people, so yeah. There would be no point for him to self-isolate somewhere else when I’m not going out” (Participant 07, Indian, female)*.

### Contact with household members

Accommodation was viewed as being important for those who are unable to isolate from their household due to the size of the house and / or the size of the household. Participants described how they would be willing to move out of the home as the amount of shared space would make isolation within the home difficult:

*“Well personally, I wouldn’t have been any choice, I think it’s the best way to prevent either him or me from getting the virus because living in the same house, it would be, uh the risk would be very high because we are sharing the same bathroom, the same kitchen, uh you know, so it would be very difficult” (Participant 07, Indian, female)*.

Among BAME communities in particular, this was considered to be a substantial problem as many within the community would live in multigenerational households:

*“That idea was a very good idea. I mean in [home town] there are areas where you have three generations living in a terraced house, grandparents, parents and the children yeah. Okay yeah now the reason why there is such a high rate of the virus here in [home town] is because of the housing here. Yeah outdated housing, and you know, because the family unit is very good, they look after each other, but because of COIVD it’s come back to haunt us big time” (Participant 12, Asian, male)*.

However, even those who had sufficient space for isolation highlighted difficulties in containing the virus and preventing the spread of viruses within the household:

*“I think personally that’s a really really good idea. Because going back to what I was saying about infection control I know how hard it must be to limit exposure if one of you’s got a virus, not just COVID, but any virus” (Participant 14, White, female)*.

### Key concerns

Participants raised a series of issues and concerns surrounding the provision of accommodation outside the home that should be addressed before such a scheme could be offered. Participants were keen to understand who should use temporary accommodation, at what stage, and for how long. Concerns were also raised among those with caring duties and responsibilities, and questions were asked regarding who would fund the scheme.

### Timing and duration

Participants wanted clarification regarding the stage at which people should move into temporary accommodation, and for how long. Participants were concerned that it would be too late to move out of the home once symptoms had presented.

*“It’s, the, to me, because all the guidance and information that we ve had is that you’re contagious before you start showing symptoms, I wouldn’t want to, because in my opinion if that is all true, you would already been exposed to it, he’d have already had it or already have it, um it just it feels like that would be too late” (Participant 15, White, female)*.

Despite concerns about leaving it too late, participants were not willing to move out of the home for long and unspecified periods of time:

“I *will be very very reluctant to go and live somewhere else. If it’s for about a week or something I don’t mind, but uh, but still yes it’s just a matter of change because we have always lived in our houses, so to go out and live somewhere else it’s quite a bit of a change” (Participant 13, Indian, male)*.

In the case of the extremely clinically vulnerable, the duration was deemed necessary to protect the family: “ *You’ve just got to get through it, and it was only like, well it could have been 12 weeks, but in a lifetime it’s not that long, really” (Participant 18, White, female)*.

### Who should use temporary accommodation

Participants raised questions about whether the intention would be for symptomatic persons or vulnerable persons to leave the home for temporary accommodation. Concerns were expressed regarding the potential of infected individuals to spread the virus should they leave the home to stay, for example, with a family member:

*“I think I would probably self-isolate too at my own house, rather than, because I might already have symptoms unknowingly, and then if I go to another household I might spread it to say, like my mum, so I think I would actually stay put” (Participant 02, Mixed White/ Black Caribbean, female)*.

In addition, concerns were also raised regarding the potential for those who are not infected to catch the virus in temporary accommodation:

*“Again I would kind of feel I would be safer at home… You go into somewhere else that I couldn’t guarantee would be as clean as I would you know, me cleaning it” (Participant 17, White, female)*.

Participants suggested schemes in which exposed workers were asked to move into temporary accommodation as a preventative measure, thus saving infection from entering the household in the first place:

*“I think almost, you’re better offering it to the workers who might go back, so like, a lot of people still worked throughout, where they couldn’t, so actually, were they the ones taking it back into their own homes, so actually would it be better targeting the workers and saying right if this happens again, if you are a key worker and you’ve got people at home, then you go to the hotel, like the NHS staff did, rather than let’s have it for people who are sick” (Participant 19, White, male)*.

For healthy individuals moving out to protect vulnerable residents, the ability to continue to work was important, and accommodation with internet access and / or within commuting distance of their work site would be necessary:

*“You know, if you were the person who was COVID free and leaving your family in the house, I don’t know which way round you suggest because if I was COVID free I’d still want to work, so it would have to be close to work*’ *(Participant 14, White, female)*.

*“If it was me going to self-isolate, for example, um, and I work from home you know, I would want, you know, I would like to be able to still have my internet and be able to carry on with my work” (Participant 01, Black African, female)*.

### Caring responsibilities

Among those who had caring responsibilities or were dependent on others, concerns were raised as to who would care for the family in their absence:

*“Um, it would be hard and difficult because you’re used to living with each other you’re reliant on each other as a family, you know, I do the shopping for the house most of the time so you know, cooking and things like that, so if I wasn’t there, or my husband wasn’t there, you know, because of the kids and all that” (Participant 04, Black British, female)*.

Participants described defined roles and responsibilities for each household member, and removal of key persons was viewed as problematic:

*I’m just wondering now what would have happened if she [participant’s wife] had the COVID 19, because she is the main person who drives the house, because she does the cooking and looks after my mum, so if she was made to go out and live somewhere else then my mum would have problems, we would have problems” (Participant 13, Indian, male)*.

Among BAME communities in particular, the need and desire to care for family members was a considerable cause for concern. Allowing others to care for their relatives was something that was only to be considered as a last resort:

*“I wouldn’t like to move out from my house, but if it is really essential then I would move, but I would try to fight it off (laughs) yeah, and I guess uh, if it happened to my mum then my mum would be the same, she wouldn’t like to live elsewhere, this was her home for the last 40 years. So because, with Indians we are very close knitted families, we tend to stick by each other, so to her it would probably do more damage going away from us than uh, and then uh, yes, than living not here” (Participant 13, Indian, male)*.

Concerns were raised about having to leave vulnerable members, potentially putting them at increased risk of exposure to the virus:

*“If I worked within the NHS and I was a key worker in that respect then possibly, but I still think just would be very difficult for me to leave the family home because of [son’s name] and again, husband and his medical condition, because he wouldn’t be able to look after my son, our son the way I would like, picking up food and medication and what not, and then he’d have to, if I wasn’t there he’d have to take the lift and sort of opening up more risk to, he’d be more in contact with people too, so I would say no in that respect” (Participant 17, White, female)*.

However, there was recognition that despite best efforts carers may contract the virus and participants had started to make tentative plans for how they would cope should this happen:

*“But that was constantly at the back of my mind like, I am going to the shops and say if I caught the virus on the handle of a trolley and then I touch my nose or my eyes and I have caught the virus now and will I have to relocate or move to my bothers house and who would care for my mother? And these were all questions at the back of my mind, but I do know my house is a 4 bedroom house and I could have self-isolate in another room and not put my mother to more risk or more harm… I would go into a separate room in the house and then sleep in the bed and then ideally move, um, not have any contact at all with my mother in the house and call my brother and ask him to intervene” (Participant 05, British Indian, male)*.

### Social and emotional support

Despite recognising the value and need for accommodation outside the home, participants struggled with the idea of having to leave the family and home:

*“If you’re forced to stay at home at least you have all of your belongings, all things that bring you comfort and people around you. But if you’re in a hotel room by yourself with just the TV and yeah, I would be so bored I think. Probably very anxious as well and quite upset. I’m such an over thinker as well so I would just be overthinking everything. But also at the same time if it meant that my partner doesn’t catch it, then I think that’s probably the main thing on my mind, if it’s temporarily a solution and hopefully that would stop the spread so I would try to look at the positive side of things, but if it was more than two weeks then yeah I really don’t know how I would deal with that” (Participant 02, Mixed White/Black Caribbean)*.

It was thought that it would be emotionally challenging to be alone and in unfamiliar surroundings:

*“I think that would be quite scary like having to do, like I mean I can’t imagine having to do this entire lockdown period by myself, like, obviously I would have to manage but there would have been a lot of different struggles with that kind of thing and I know people who have done it have been lonely and it would have taken a while to adapt, it would be really difficult” (Participant 10, Asian British, female)*.

Participants highlighted the need for facilities to enable them to continue to communicate with their friends and family throughout:

*“I have a lot of, all my social stuff is now online, so my theatre group, we rehearse online, we have various support groups and stuff, so for me it would be very important to still be able to have that” (Participant 01, Black African, female)*.

### Essential requirements

Whilst all participants reported requiring only the basics, further detail regarding food, washing and cleaning facilities were needed:

*“I think a room with internet, and uh a bathroom and then just an understanding of how the uh meal system will work” (Participant 08, Indian, male)*.

Food in particular was a key concern:

*“Um, to be able to cook my own food, for me food is very important to me, it is to everybody, but not everyone has the kind of attention to what they eat, I don’t eat meat, um, so um, you know, I eat fish but I, yeah I like to have my own space to cook my food” (Participant 01, Black African, female)*.

Indeed, there were reports of food related complaints from other locations within which this system is widely implemented:

*“Well in the beginning they [residents in isolation facilities in [country]] were really complaining about the food that they were getting in the centres… and yeah after one or three weeks, I mean, I guess maybe they changed the types of food they were getting” (Participant 07, Indian, female)*.

Those who had used accommodation to avoid transmitting the virus to vulnerable members of the household described how they had had to work hard to ensure food and cooking facilities were available:

*“He had local chip shops offering to cook him food, especially in the early days when we didn’t really know, we hadn’t really found our routine, so like the local fish and chip shop were feeding him, to be fair the people who run the hotel were feeding him, because they live on site, he had work colleagues bringing him plates of food, people dropping him food off, and then we kind of found routine, somebody gave him a microwave, somebody else gave him a fridge, somebody else gave him a toaster. It was a real community effort. Yeah after about 3 or 4 weeks he fell into a routine and he could cook himself stuff so it wasn’t so bad” (Participant 18, White, female)*.

Those in temporary accommodation could also provide tangible support for vulnerable members of the family at home:

*“So for that rocky stage when people were struggling [to secure priority slots], yeah I had a little servant on the outside” (Participant 18, White, female)*.

Participants also described a need for outside space to maintain physical and emotional health:

*“I’d need to be able to get outside, to have, like here I have a garden here, so it’s just to be able to, you know, even when it’s raining I walk out to the garden just to get some air” (Participant 01, Black African, female)*

Indeed, outside space for physical activity was considered invaluable to those who had moved out of the home:

*“He runs. A lot. An awful lot. So yeah that is how he coped. Yeah, and like initially we thought it was going to be a lot harder the lockdown, so the first week he thought I’m just going to run when I can because we thought exercise was going to be stopped. So he kind of hit the 50 mile a week mark, and then it didn’t stop, so he just kept that up really. Just running every day” (Participant 18, White female)*.

### Funding

Participants were concerned about costs associated with temporary accommodation. Participants were unable to cover the costs themselves, and the one participant whose husband had used accommodation to prevent transmission of the virus to her vulnerable daughter described how it was only possible because it was free of charge. Although the costs were later covered by the National Health Service (NHS), she described how it would not have been possible to pay for accommodation without the goodwill of the community:

*“He was really lucky because I know a lot of NHS workers had to wait to move out because NHS trusts and health boards took a while to get their system working, but one of the local hotels, because we live in quite a small area, one of the local hotels offered free rooms, so he was actually able to move out straight a way, on that very first Monday he was out. So, yeah. And the health board did pay in the end, but it was right at the end that they decided they were going to pay for it, but the hotel would have given him free room for like 10 weeks, 10, 11 weeks” (Participant 18, White, female)*.

Despite the lifesaving potential of the scheme, many were unconvinced that it would be funded by the current government:

*“I mean, in all honesty I would be like incredibly surprised if um that was like, if this current government were offering that to people” (Participant 03, British Asian/Pakistani)*.

*“Yeah. So that is a very splendid idea if that was possible, but economically it’s not viable is it? It’s a good option but economically I don’t think this government would go for it anyway. But yeah it’s a very good system that if it was in place. Yeah.” (Participant 12, Asian, male)*.

## Discussion

### Summary of findings

To our knowledge, this is the first study to have explored issues surrounding the option of accommodation to prevent transmission of the virus in vulnerable households. This work reveals that the offer of accommodation to protect vulnerable households is viewed positively by many people who feel their household is at risk. Data collected from both survey and interview participants highlighted concerns regarding the spread of the virus within the household, and a need for solutions to prevent this. Interviews provided insight into populations who would be likely to accept and benefit from the offer; and it was suggested that those who are vulnerable, are likely to be exposed to the virus, and who are unable to isolate within the home would benefit most. Participants who met one or more of these criteria appeared very willing to accept the offer of accommodation compared with those who consider themselves or their household not to be vulnerable, were not employed in public facing occupations, and/or had capacity to isolate within the home. Crucially, and in line with existing research (12), those from BAME and low income communities were considered to be more exposed and less able to isolate than those from high income backgrounds.

Participants questioned who should use temporary accommodation and at what stage, with legitimate concerns being raised regarding the utility of isolating outside the home once symptoms are present.

In locations in which accommodation has successfully been used to support self-isolation outside the home, it is often the symptomatic persons who are offered accommodation for isolation (19, 20). In the UK, NHS staff who are living with vulnerable family members have been offered accommodation to protect the family whilst allowing those not at high risk to continue to work (28). Our study suggests that both approaches could be feasible and acceptable to high risk audiences, but the offer of accommodation must be timely, and appropriate infection control measures must be in place.

Different households will have different requirements – there is no ‘one size fits all’. However, as lockdown restrictions are lifted, and test, trace and isolate becomes a key strategy in controlling the virus, making support available to allow certain individuals to isolate safely could make a potentially valuable contribution to reducing transmission, morbidity and mortality.

### Implications of this study

This study revealed some important issues that would need to be addressed to ensure the acceptability and feasibility of any offer of accommodation for those who need it. Drawing together findings from the survey and interviews we consider below some of the options available, key concerns associated with isolating outside the home, and ways in which these may be mitigated.

In locations in which accommodation is provided, it is the individual with the virus who would isolate outside the home in order to protect vulnerable household members (20). Participants were concerned that, by the time symptoms were evident, transmission of the virus to other household members would already have occurred. However, with a test, trace and isolate system firmly in place, it would be possible for those who have been in contact with virus to be offered accommodation to quarantine before symptoms emerge. Indeed, individuals who are informed that they have been in contact with the virus may not be willing to return to their homes to await test results if they are living with vulnerable relatives. The offer of accommodation for individuals in this situation could be highly effective.

A second option for utilising accommodation to prevent transmission of the virus within the household involves moving vulnerable people out of the home should household members become symptomatic. Although this was seen as a viable option, again, there were concerns that it would be too late to make use of temporary accommodation at the stage at which infected persons are showing symptoms. There is however, emerging evidence to suggest that viral load is associated with disease severity (29), and initial viral load is likely to be a contributing factor (30). Interventions aiming to reduce exposure to the virus in the home have been successful (31, 32). However, it is not easy to avoid contact with infected individuals, and more needs to be done to support vulnerable people (33). In particular, vulnerable individuals living in large households may be at risk of exposure to a high viral load from multiple sources if support is not available. In such situations, offering accommodation to vulnerable individuals, with appropriate care and support, could substantially reduce their exposure to the virus.

Participants recognised the significant practical and emotional challenges associated with utilising accommodation to prevent transmission of the virus, and it is critical that those who are in quarantine or isolating outside (and inside) the home are adequately supported. Both practical (e.g., food) and emotional support will be required, for example through community support networks, similar to those that were established at the start of the pandemic. Participants also raised critically important concerns about exposure to infection in temporary accommodation that must be addressed. Strategies must be put in place to ensure that those in temporary accommodation are not exposed, or exposing others, to the virus.

There were concerns over who would fund accommodation, and indeed, it would not be cost effective to provide accommodation for all populations. However, we suggest that offering accommodation in a targeted way to those who are vulnerable, exposed to the virus, and/or unable to quarantine or isolate safely within their home would reduce these costs, and could even lead to a potential reduction in healthcare costs if the number of vulnerable individuals exposed to the virus is reduced.

### Limitations

The main limitation associated with this work is the extent to which the views of our sample are representative of the UK population. Our recruitment for phase one occurred via a mailing list of individuals who had previously used and provided feedback on a website aiming to reduce infection within the home. This is therefore likely to be a group of individuals who are highly motivated to engage in infection control behaviours and their views may not be representative of the wider population. However, those who took part in the interview phase of the study had not shown any prior interest in infection control practices. Whilst every effort was made to recruit a diverse sample of participants for interviews, our primary use of social media to recruit participants may have resulted in individuals with very relevant voices being excluded. For example, those from non-English speaking communities, those without internet access, and those without social media would have been missed. Despite attempts to recruit participants though existing networks with community group leaders, engagement through these networks was minimal and could not be pursued further due to the need for timely completion of this initial study.

The rapidly changing nature of the pandemic and government advice limits the interpretation of our findings. As perceptions of risk within and outside the home change, the acceptability of accommodation to prevent transmission of the virus in the home may also shift. Our findings must be interpreted with this in mind.

### Conclusions

Within-household transmission is likely to be a leading cause of morbidity and mortality as we move out of lockdown (31) and we present just some of the ways in which accommodation may be viewed and utilised. We recognise the complexities associated with these options, and acknowledge that different households will require very different provisions. However, we suggest that offering accommodation to vulnerable households following a potential exposure to the virus, or during the early stages of an outbreak within the home could be acceptable and feasible.

## Data Availability

The datasets used and/or analysed during the current study are available from the corresponding author on reasonable request.

## Declarations

### Ethics approval and consent to participate

Ethical approval for Phase 1 was provided by Southampton Research Ethics Committee (56445). All survey participants consented online to take part in the study.

Ethical approval for Phase 2 was provided by the NHS Health Research Authority London – Queen Square Research Ethics Committee (20/HRA/2549). All interview participants verbally consented to take part in the study.

### Consent for publication

All participants provided verbal or written consent for data to be included in publications.

### Authors’ contributions

Conceived the study: LY, SD, JH

Study design: LY, SD, JH

Analysed the data: SD, KM, RdG

Interpreted the data: All authors

Drafted the manuscript: SD

Reviewed the manuscript and approved content: All authors

Met authorship criteria: All authors

## Acknowledgements

Lucy Yardley is an NIHR Senior Investigator and her research programme is partly supported by NIHR Applied Research Collaboration (ARC)-West, NIHR Health Protection Research Unit (HPRU) in Behavioural Science and Evaluation, and the NIHR Southampton Biomedical Research Centre (BRC). Jeremy Horwood is partly supported by NIHR Applied Research Collaboration (ARC)-West, and NIHR Health Protection Research Unit (HPRU) for Behavioural Science and Evaluation at the University of Bristol. Rachel de Garang is a BME Engagement Worker for the Voice & Influence Partnership at The Care Forum.

